# Predictors of Time to Start of Trophic Feeding in Preterm Neonates Admitted to Neonatal Intensive Care Unit of Adama Hospital Medical College, Ethiopia: A Retrospective Cohort Study

**DOI:** 10.64898/2026.08.26.26361481

**Authors:** Bonso Misha, Godana Arero Dassie, Ibrahim Mohammed

**Author notes:** Corresponding author. Godana Arero Dassie.

## Abstract

**Background:** Early trophic feeding promotes gut maturation, feeding tolerance, and growth in preterm neonates. However, delays remain common despite recommendations for initiation within 24 hours of birth, especially in resource-limited settings. Evidence on feeding initiation timing and predictors among Ethiopian preterm neonates is limited.

**Objective:** To determine time to trophic feeding initiation and identify predictors among preterm neonates admitted to Adama Hospital Medical College, Ethiopia.

**Methods:** A hospital-based retrospective cohort study was performed on 436 randomly chosen preterm neonates admitted to NICU. Data extraction was performed using a structured checklist. Time to trophic feeding initiation was analyzed using Kaplan–Meier estimates, log-rank tests, and bivariable and multivariable Cox regression models . Adjusted hazard ratios with 95% CIs were reported.

**Results:** The sample comprised 416 preterm neonates, of whom 311 (74.8%) started trophic feeding during follow-up, and 105 (25.2%) were censored. The rate of initiation of trophic feeding was 1.92 per 100 person-hours (95% CI 1.72 to 2.15). Median time to initiation was 42 hours (interquartile range 24 to 50). Independent predictors of feeding initiation were determined by multivariable analysis and included gestational age, birth weight, maternal anaemia, respiratory distress syndrome and necrotising enterocolitis. Neonates born at 34–36 weeks had earlier initiation than those born at <34 weeks (AHR 1.39; 95 % CI 1.09 to 1.78). Similarly, neonates with a birth weight of ≥1500 g had an earlier initiation than those with a birth weight of <1500 g (AHR 1.41; 95% CI 1.04 to 1.91). Delayed initiation was associated with maternal anaemia (AHR 0.70; 95% CI 0.51–0.95), respiratory distress syndrome (AHR 0.67; 95% CI 0.51–0.88) and necrotising enterocolitis (AHR 0.48; 95% CI 0.33–0.69).

**Conclusions:** Delayed trophic feeding remains common among preterm neonates. Standardized feeding protocols, strengthened maternal care, and individualized nutrition strategies are needed to improve neonatal outcomes in study area.

**Key messages:** *What is already known about the subject?:* ✓ ◻Early trophic feeding aids in intestinal maturation, improves feeding tolerance and reduces the complications of long-term nutritional deprivation in preterm infants. ✓ ◻International recommendations advocate that minimal enteral feeding should be started within the first 24 hours of life whenever clinically feasible. ✓ Delayed trophic feeding is still a common practice in neonatal intensive care units particularly in low resource settings. However, there is scanty evidence on determinants of delayed initiation in Ethiopia.

## 1. Introduction

The World Health Organization (WHO) describes preterm birth as giving birth before 37 weeks of pregnancy are completed. Preterm birth is still a major global public health challenge and the leading cause of neonatal morbidity and mortality. An estimated 13.4 million infants were born preterm in 2020, accounting for 9.9% of all live births, with the burden heavily concentrated in South Asia and sub-Saharan Africa [1,3]. Optimizing early nutritional management remains critical despite advances in neonatal intensive care because preterm neonates are at increased risk of growth failure, severe complications and death due to gastrointestinal immaturity, limited nutrient reserves and clinical instability [4,23].

Trophic feeding, or minimal enteral nutrition, is the infusion of small amounts of enteral formula, preferably colostrum or mother’s own breast milk, primarily for the purpose of stimulating gastrointestinal development rather than full nutritional support [5,9]. International recommendations for neonatal nutrition suggest that trophic feeding should be started early, within the first 24 h of life in clinically stable preterm infants and advanced progressively according to tolerance and clinical condition [15,16]. Early trophic feeding promotes intestinal motility, stimulates the release of gastrointestinal hormones, maintains the integrity of the mucosa, facilitates maturation of the gut, speeds the transition to full enteral feeding, and reduces the dependence on prolonged parenteral nutrition [9,11]. Systematic reviews have shown that early enteral feeding is safe and may improve feeding tolerance without increasing the risk of necrotizing enterocolitis (NEC) when appropriate feeding strategies are used [5–8].

Early trophic feeding is recommended as an essential part of neonatal care; however, implementation varies substantially among neonatal intensive care units (NICUs). Differences in feeding protocols, access to human milk, practices of healthcare providers, severity of neonatal illness, and health system resources influence variation in feeding initiation practices [17,30,32]. Therefore, many preterm infants have delayed introduction of enteral nutrition despite evidence of the safety and physiological benefits of early minimal enteral feeding.

Delayed introduction of trophic feeds may negatively impact neonatal outcomes through delayed gastrointestinal maturation, prolonged hospitalization, feeding intolerance, impaired postnatal growth, increased dependence on parenteral nutrition, and increased risk of neonatal complications including sepsis and NEC [6,10,17]. Growth restriction after birth is still common in preterm infants and partly associated with inadequate nutrition during their hospital stay [18,23]. Prior research indicates that a significant number of very preterm and very low birth weight infants are not fed enterally during the first 24–48 hours of life, highlighting ongoing disparities between evidence-based recommendations and routine clinical practice [30,31].

In Ethiopia the problem of babies being born early and the issues that come with it are really big deals. A lot of babies die because they were born early [1,3]. Although national neonatal care initiatives and improvements in NICU services emphasize early enteral nutrition and evidence-based feeding practices [25,27], delayed initiation of trophic feeds is still common. However, in Ethiopia, only some of the preterm neonates received trophic feeding in the first 24 hours, with median time to initiation ranging between 40.8 and 41.7 hours after birth[15,31]. The results show that we can do a lot to make sure newborn babies get the food they need by doing things the way every time and by coming up with better plans to make it all work.

Initiation of trophic feeding is dependent on various maternal and neonatal factors. Delayed enteral feeding in preterm infants has been previously reported to be associated with lower gestational age, very low birth weight, respiratory distress syndrome, low APGAR scores, maternal complications, and gastrointestinal disorders [4,15,23,31,32]. However, most Ethiopian studies have focused on the proportion of infants receiving early feeding, rather than using time-to-event approaches to assess the patterns and predictors of feeding initiation. Also, there are limited data from NICUs outside major urban referral centres.

Therefore, this retrospective cohort study was conducted to determine the time to initiation of trophic feeding and to identify independent predictors among preterm neonates admitted to the Neonatal Intensive Care Unit of Adama Hospital Medical College, Southeast Ethiopia, using Kaplan–Meier survival analysis and multivariable Cox proportional hazards regression. The findings offer evidence to strengthen neonatal feeding protocols, improve nutritional care practices, and support policy development for the management of preterm infants in resource-limited settings.

## 3. Methodology

### 3.1 Study Design and Reporting Guideline

A hospital based retrospective cohort study was carried out in Neonatal Intensive Care Unit (NICU) of Adama hospital medical college, south east Ethiopia to find out time to initiation of trophic feeding and its predictors among preterm neonates. The retrospective cohort design was deemed appropriate as it allowed the evaluation of time-to-event outcomes using routinely collected clinical data while reducing the cost and time associated with prospective follow-up. The study was reported in accordance with the Strengthening the Reporting of Observational Studies in Epidemiology (STROBE) statement for cohort studies to ensure transparent and complete reporting of the study design, participant selection, data collection, statistical analyses and interpretation of findings.

### 3.2 Study setting and period

The study was conducted at the Neonatal Intensive Care Unit (NICU) of Adama Hospital Medical College, a tertiary teaching and referral hospital found in Adama City, Oromia Regional State, about 99 km southeast of Addis Ababa, Ethiopia. The hospital offers a full referral service in maternal, neonatal and paediatric care to a population of more than six million people in central and south eastern Ethiopia.

The NICU admits critically ill and high-risk neonates delivered in the hospital as well as referrals from the surrounding hospitals and health centers. The unit is divided into five functional sections: a front stabilization unit (three beds), a critical care unit (17 beds), a preterm unit (eight beds), a kangaroo mother care unit (seven beds) and a stable neonatal care unit (15 beds). The NICU contains five incubators, ten radiant warmers, phototherapy units, infusion pumps, and standard neonatal monitoring equipment, as well as continuous positive airway pressure devices. The clinical care is provided by a multidisciplinary team consisting of one neonatologist, six pediatricians, ten neonatal nurses, fifteen Bachelor of Science nurses and one clinical nurse.

Approximately 600 premature babies are admitted each year. Data were collected from July 1, 2024 to July 30, 2024. All medical records of preterm neonates admitted between January 1, 2023 and December 30, 2023 were reviewed.

### 3.3 Population of Study

The source population comprised all medical records of preterm neonates admitted to NICU of Adama Hospital Medical College during the study period. The study population was selected randomly from medical records of preterm neonates who met the pre-set inclusion criteria. The unit of analysis was a single preterm neonate.

### 3.4 Criteria for Eligibility

We included medical records of neonates born before 37 completed weeks of gestation and admitted to the NICU during the study period. Records were excluded if they were missing or inconsistent with essential information needed to determine the outcome or key explanatory variables, such as gestational age, birth date and time, APGAR score, birth weight, or documented time of initiation of trophic feeding. Neonates who had already been started on enteral feeding before NICU admission were excluded since the exact timing of initiation of trophic feeding could not be reliably determined. Simple random sampling was used to give each and every eligible record an equal chance of being selected so as to avoid selection bias.

### 3.5 Sample Size Estimation

Sample size was determined using the Schoenfeld method for Cox proportional hazards regression on predictors of initiation of trophic feeding among preterm neonates in Ethiopia as reported in previous studies. This calculation was based on a 5% significance level (two sided), 80% power, 90.9% event probability and 10% allowance for incomplete records. Sample size estimates were generated for several clinical relevant predictors including gestational age, APGAR score, respiratory distress syndrome, perinatal asphyxia, hemodynamic instability, mode of delivery and place of delivery. The largest sample size was for gestational age as a predictor, and hence the final required sample was 436 preterm neonates. Sample size was determined using STATA version 17.

### 3.6 Method of Sampling

Simple random sampling technique was used. First, all preterm admissions recorded in the NICU registration log during the study period were identified, yielding a sampling frame of 612 eligible medical registration numbers. Then, a sample of 436 records was selected by a computer-generated random number sequence. Individual medical charts were retrieved from the hospital archive using selected registration numbers. This probability sampling method minimized selection bias and improved the representativeness of the study population.

### 3.7 Variable of the Study

The primary outcome measure was the time to initiation of trophic feeding, which was defined as the number of hours from birth to the first documented administration of trophic enteral feeding. We defined the event (event = 1) as neonates who initiated trophic feeding during hospitalization. Neonates who died, transferred, referred, discharged before trophic feeding or remained without documented trophic feeding at the end of follow-up were considered as censored observations (event = 0).

Potential explanatory variables included maternal sociodemographic characteristics, obstetric factors, neonatal characteristics and neonatal clinical conditions. Maternal variables included age, place of residence, attendance of antenatal care, parity, multiple pregnancy, pregnancy complication, anemia, diabetes mellitus, human immunodeficiency virus infection, pre-eclampsia, eclampsia, place of delivery and mode of delivery. The neonatal variables included sex, gestational age, birth weight, weight-for-gestational-age category, APGAR score, age at admission, respiratory distress syndrome, neonatal sepsis, necrotizing enterocolitis, perinatal asphyxia, jaundice, hypothermia, hypoglycemia and neonatal anemia.

### 3.8 Tool and Procedure for Data Collection

Data were abstracted using a structured, pretested data abstraction checklist developed after review of published literature, neonatal nutrition guidelines and standard NICU medical records. The checklist recorded maternal characteristics, obstetric history, neonatal demographic and clinical characteristics, laboratory findings, feeding practices, and hospitalization outcomes. Information was extracted from delivery records, admission notes, physician’s progress notes, nursing records, feeding charts and discharge summaries.

Data were collected by three trained BSc nurses under the supervision of one experienced neonatal nurse. All data collectors were trained using a standard protocol on study objectives, variable definitions, abstraction procedures, confidentiality and data quality assurance before data collection. The principal investigator supervised the whole study.

### 3.9 Data Quality Assurance

To ensure the quality of data, the abstraction checklist was pretested on approximately 5% of the calculated sample in Mojo General Hospital and this was not included in the main study. Where necessary changes were made in light of the pilot findings. Data collectors and supervisors were trained in a standardized way before data collection. The supervisor and principal investigator reviewed abstraction forms on a daily basis for completeness, consistency, and accuracy. Data were double checked before data entry. Range and logical checks were done during data entry and discrepancies were resolved by re-examination of the original medical records. Complete case analysis was done as those with missing data regarding the outcome or key predictors were not included in the selection of participants.

### 3.10 Statistics Analysis

Data were entered into EpiData version 4.6 with programmed validation checks and exported to STATA version 17 for cleaning and statistical analysis. Continuous variables were presented as means and standard deviations or medians and interquartile ranges depending on their distribution. Categorical variables were presented as frequencies and percentages.

Survival time was recorded from birth until the start of trophic feeding and expressed in hours. Cumulative probability of not starting trophic feeds over time was estimated using Kaplan-Meier survival analysis, and differences between survival curves were determined using the log-rank test . Incidence rates were calculated as per person-hours of follow-up.

Bivariable Cox proportional hazards regression was performed to identify candidate predictors, and variables with a p-value <0.25 were entered into the multivariable Cox proportional hazards model to control for potential confounding. Adjusted hazard ratios (AHRs) with 95% confidence intervals were computed. A p-value < 0.05 was considered statistically significant.

No indication of problematic multicollinearity was observed, and multicollinearity between the explanatory variables was tested using variance inflation factors before fitting the model. Potential interaction between clinically relevant variables where explored, but no statistically significant interaction terms were retained in final model. The proportional hazards assumption was tested with Schoenfeld residuals and no violation of the assumption was found (global test p = 0.5253). The overall adequacy of the model was checked by Cox – Snell residual plots and the fit of the model was found to be satisfactory. Sensitivity analyses of excluded and included records showed no meaningful differences in major baseline characteristics.

### 3.11 Reduction of Bias

Several steps were taken to reduce possible sources of bias. We used simple random sampling of all eligible medical records from a complete NICU admission registry to minimize selection bias. We minimized information bias by using standardized operational definitions, a pretested abstraction checklist, trained data collectors, and supervisor verification of extracted information. We used physician-diagnosed clinical diagnoses and standardized hospital records to reduce misclassification bias. After careful review of potential variables to account for confounding, multivariable Cox proportional hazards regression analysis was performed.

## 3. Results

### 3.1 Participant selection and baseline characteristics

A total of 436 medical records of preterm neonates admitted to the Neonatal Intensive Care Unit (NICU) at Adama Hospital Medical College during the study period were randomly selected for review. Of these, 20 records were excluded because of incomplete or insufficient information, leaving 416 neonates who fulfilled the eligibility criteria and were included in the final analysis. Thus, the overall chart completeness rate was 95.4%. The participant selection process is presented in Supplementary Figure S1. The study cohort contributed a total of 16,123 person-hours of observation for the time-to-event analysis.

Among the 416 preterm neonates, 216 (51.9%) were female and 200 (48.1%) were male. The median maternal age was 28 years (IQR, 24–32 years), with more than half of the mothers aged 25–34 years (225, 54.1%). Most mothers resided in Adama (270, 64.9%), whereas 146 (35.1%) were referred from surrounding districts. Overall, 245 (58.9%) neonates were born at 34–36 weeks of gestation and 171 (41.1%) were born at 28–33 weeks. The mean birth weight was 1,936.3 ± 507.7 g, with 332 (79.8%) neonates weighing ≥1500 g and 84 (20.2%) weighing <1500 g. The mean first-minute and fifth-minute APGAR scores were 5.3 ± 1.7 and 7.0 ± 2.1, respectively; 290 (69.7%) neonates had a first-minute APGAR score <7. The maternal and neonatal baseline characteristics are summarized in Table 1.

**Table 1.** Baseline maternal and neonatal characteristics of preterm neonates admitted to the NICU (n=416) summarizes the maternal and neonatal demographic and clinical characteristics of the 416 preterm neonates included in the analysis. APGAR, Appearance, Pulse, Grimace, Activity, and Respiration; NICU, Neonatal Intensive Care Unit.

| Characteristic | Frequency (%) |
| --- | --- |
| Maternal age (years) |  |
| <25 | 120 (28.8) |
| 25–34 | 225 (54.1) |
| $\geq 35$ | 71 (17.1) |
| Maternal residence |  |
| Adama | 270 (64.9) |
| Outside Adama | 146 (35.1) |
| Sex of neonate |  |
| Male | 200 (48.1) |
| Female | 216 (51.9) |
| Gestational age at birth |  |
| 28–33 weeks | 171 (41.1) |
| 34–36 weeks | 245 (58.9) |
| Birth weight |  |
| <1500 g | 84 (20.2) |
| $\geq 1500$ g | 332 (79.8) |
| First-minute APGAR score |  |
| <7 | 290 (69.7) |
| $\geq 7$ | 126 (30.3) |

### 3.2 Maternal medical conditions and neonatal clinical characteristics

Maternal medical conditions were common among the study participants. Maternal anaemia was the most frequently documented condition, affecting 110 (26.4%) mothers, followed by eclampsia in 95 (22.8%), hypertension in 58 (13.9%), pre-eclampsia in 54 (13.0%), and diabetes mellitus in 50 (12.0%). Syphilis was documented in 36 (8.7%) mothers, while postnatal depression and HIV infection were recorded in 23 (5.5%) and 10 (2.4%) mothers, respectively.

Several neonatal clinical complications were also documented during NICU admission. Respiratory distress syndrome was the most frequently recorded neonatal condition, occurring in 114 (27.4%) neonates, followed closely by hypoglycaemia in 113 (27.2%) and hypothermia in 92 (22.1%). Neonatal anaemia was documented in 63 (15.1%) neonates and necrotizing enterocolitis in 48 (11.5%). Other conditions included neonatal jaundice (9.9%), sepsis (9.4%), meningitis (8.9%), perinatal asphyxia (7.9%), and congenital malformations (5.5%). Maternal conditions and neonatal clinical complications considered in subsequent survival and regression analyses are presented together in Table 2.

**Table 2.** Maternal medical conditions and neonatal clinical conditions among preterm neonates (n=416) presents the distribution of documented maternal medical conditions and neonatal clinical complications among the study participants. Conditions shown in the table were evaluated as potential determinants of time to trophic feeding initiation.

| Condition | Frequency (%) |
| --- | --- |
| Maternal medical conditions |  |
| Anaemia | 110 (26.4) |
| Eclampsia | 95 (22.8) |
| Hypertension | 58 (13.9) |
| Pre-eclampsia | 54 (13.0) |
| Diabetes mellitus | 50 (12.0) |
| Syphilis | 36 (8.7) |
| Postnatal depression | 23 (5.5) |
| HIV infection | 10 (2.4) |
| Neonatal clinical conditions | Frequency (%) |
| Respiratory distress syndrome | 114 (27.4) |
| Hypoglycaemia | 113 (27.2) |
| Hypothermia | 92 (22.1) |
| Neonatal anaemia | 63 (15.1) |
| Necrotizing enterocolitis | 48 (11.5) |
| Neonatal jaundice | 41 (9.9) |
| Sepsis | 39 (9.4) |
| Meningitis | 37 (8.9) |
| Perinatal asphyxia | 33 (7.9) |
| Congenital malformation | 23 (5.5) |

### 3.3 Time to initiation of trophic feeding

During follow-up, 311 (74.8%) of the 416 preterm neonates initiated trophic feeding, whereas 105 (25.2%) were censored before feeding initiation because of death, referral, or discharge. The total observation time was 16,123 person-hours, corresponding to an overall trophic feeding initiation rate of 1.92 per 100 person-hours (95% CI, 1.72–2.15). The median time from birth to initiation of trophic feeding was 42 hours (IQR, 24–50 hours).

The cumulative incidence of trophic feeding initiation increased progressively during the first two days of life. Only 86 (20.7%) neonates had initiated trophic feeding within the first 24 hours, increasing to 220 (52.9%) by 48 hours. Thus, nearly half of the cohort had not commenced trophic feeding by 48 hours of life. The overall time-to-event distribution is summarized in Table 3 and illustrated by the Kaplan–Meier curve in Figure 2. The findings demonstrate that trophic feeding initiation was generally gradual, with a substantial proportion of preterm neonates experiencing initiation beyond the first 24–48 hours of life ( Table 3, Figure 2).

**Figure 1.**
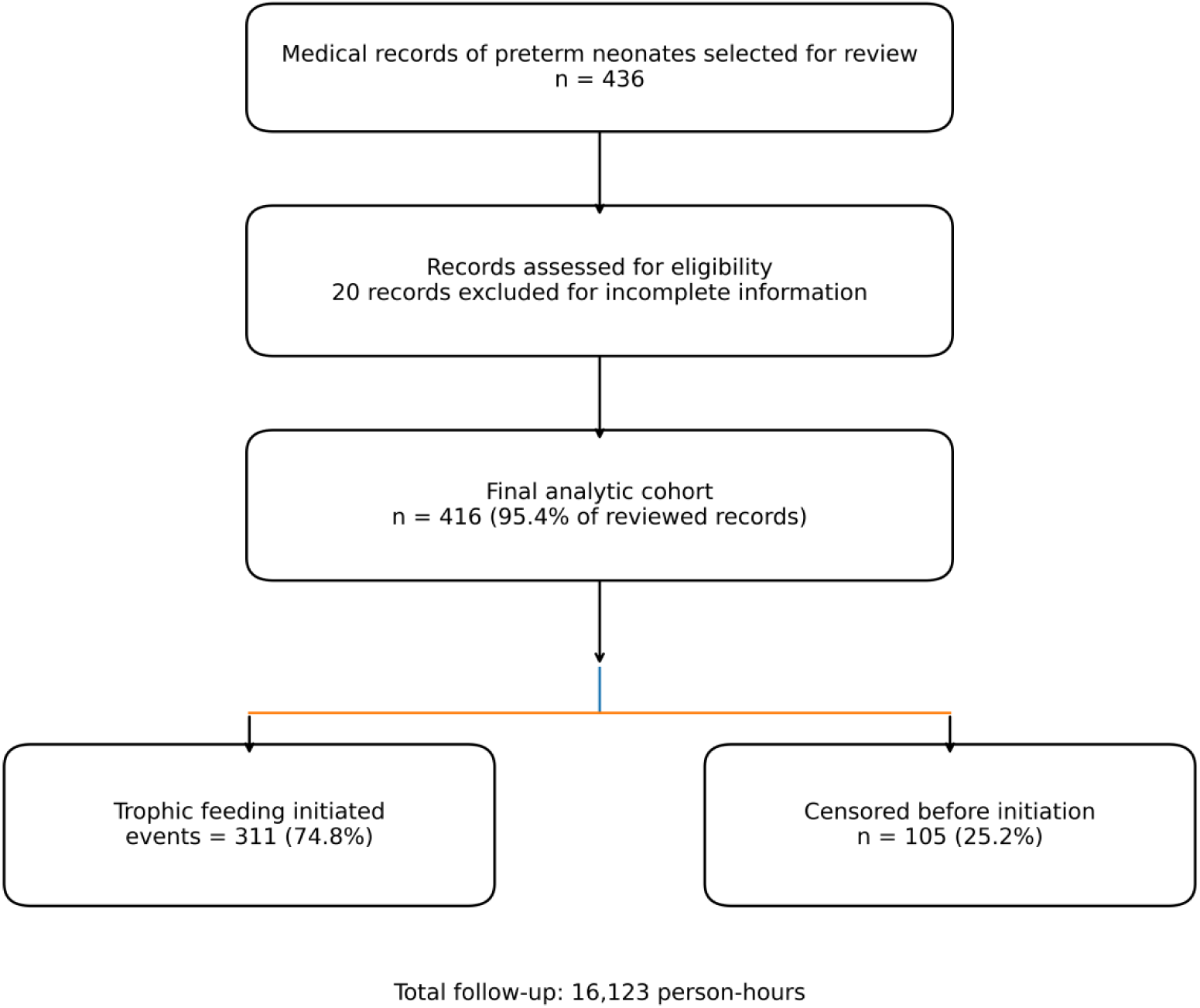
Flow diagram of selection and inclusion of preterm nenates. provides the participant selection and exclusion process, while the complete analytical dataset, variable definitions, and additional model diagnostics should be provided in the Supplementary Materials to facilitate reproducibility and transparent reporting.

**Figure 2.**
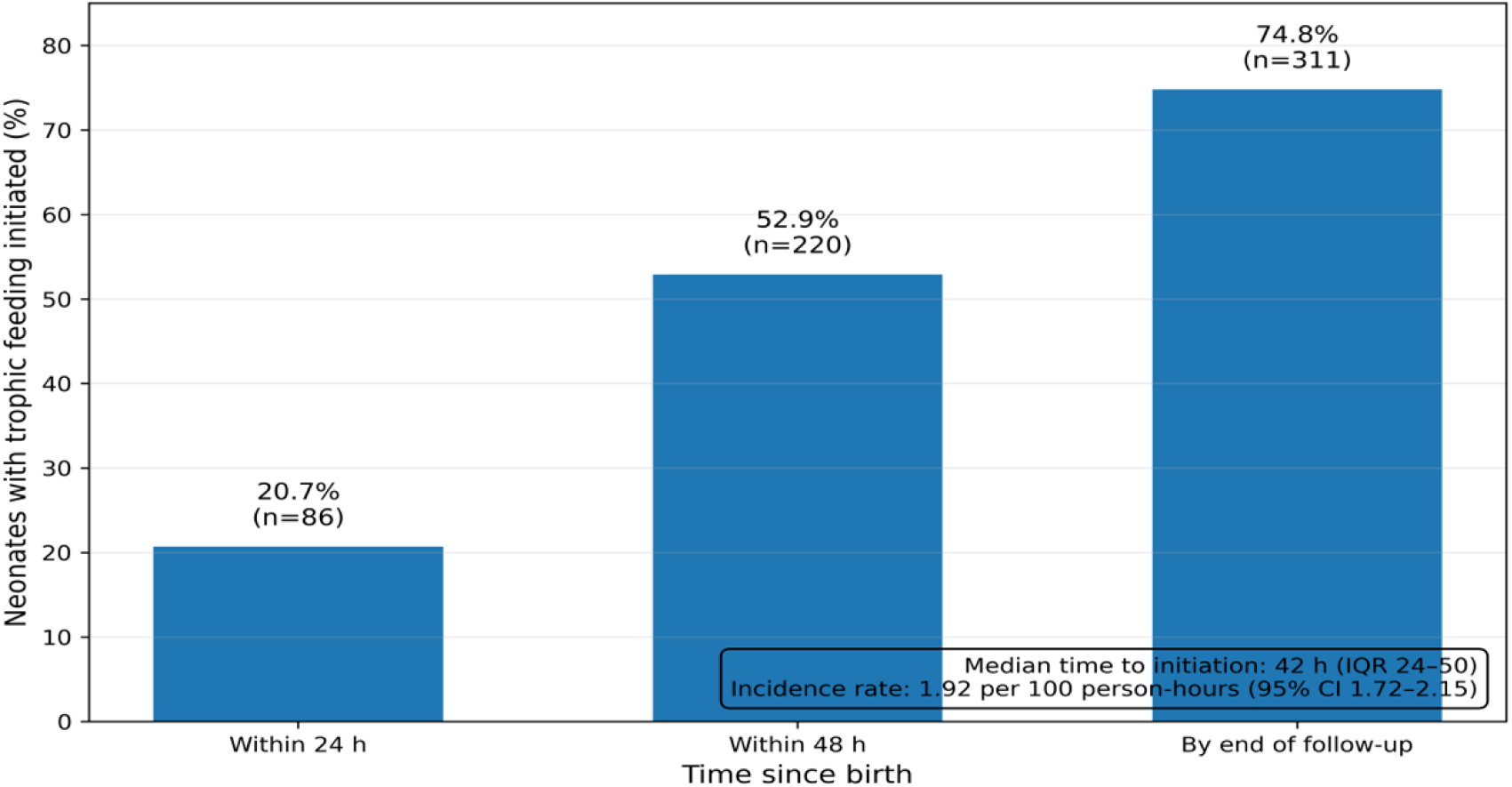
Cummulative milestones of trophic feeding initiations. The bar graph shows the cumulative percentage of neonates initiating trophic feeding after birth, rising from 20.7% ($n = 86$) within 24 hours to 52.9% ($n = 220$) within 48 hours, and reaching 74.8% ($n = 311$) by the end of follow-up. Overall, the median time to initiation was 42 hours ($\text{IQR } 24\text{–}50$), with an incidence rate of 1.92 per 100 person-hours ($95\%\text{ CI } 1.72\text{–}2.15$).

**Table 3.**
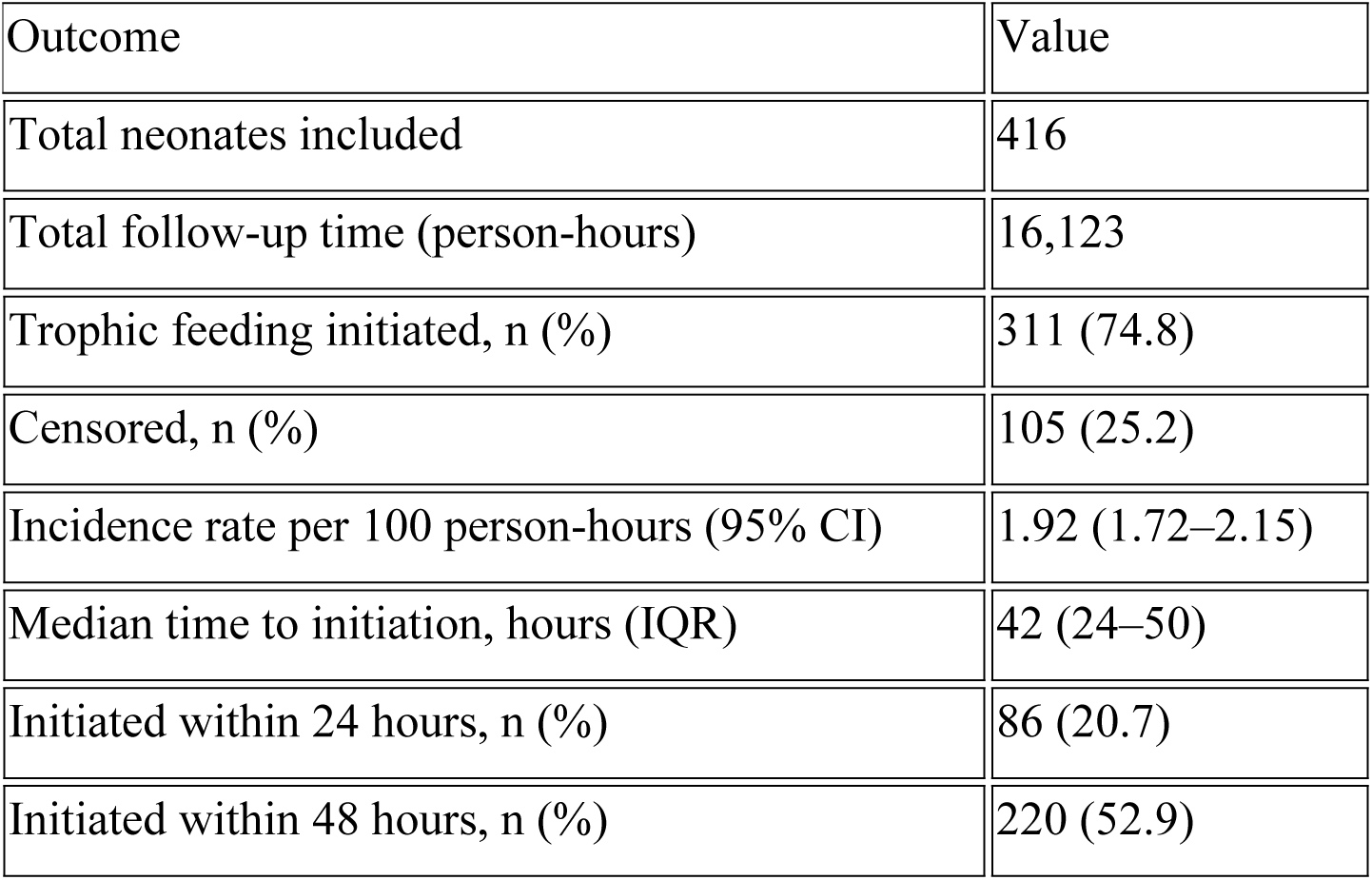
Time-to-event outcomes for trophic feeding initiation among preterm neonates (n=416) summarizes the overall time-to-event experience of the cohort. The event was defined as initiation of enteral trophic feeding. Neonates who died, were referred, or were discharged before initiation of trophic feeding were treated as censored observations.

### 4 Kaplan–Meier analysis by maternal and neonatal characteristics

Kaplan–Meier survival analysis was used to compare time to trophic feeding initiation across maternal and neonatal characteristics. Statistically significant differences were observed according to gestational age, birth weight, maternal anaemia, maternal eclampsia, respiratory distress syndrome, and necrotizing enterocolitis (Figure 3A–F).

**Figure 3.**
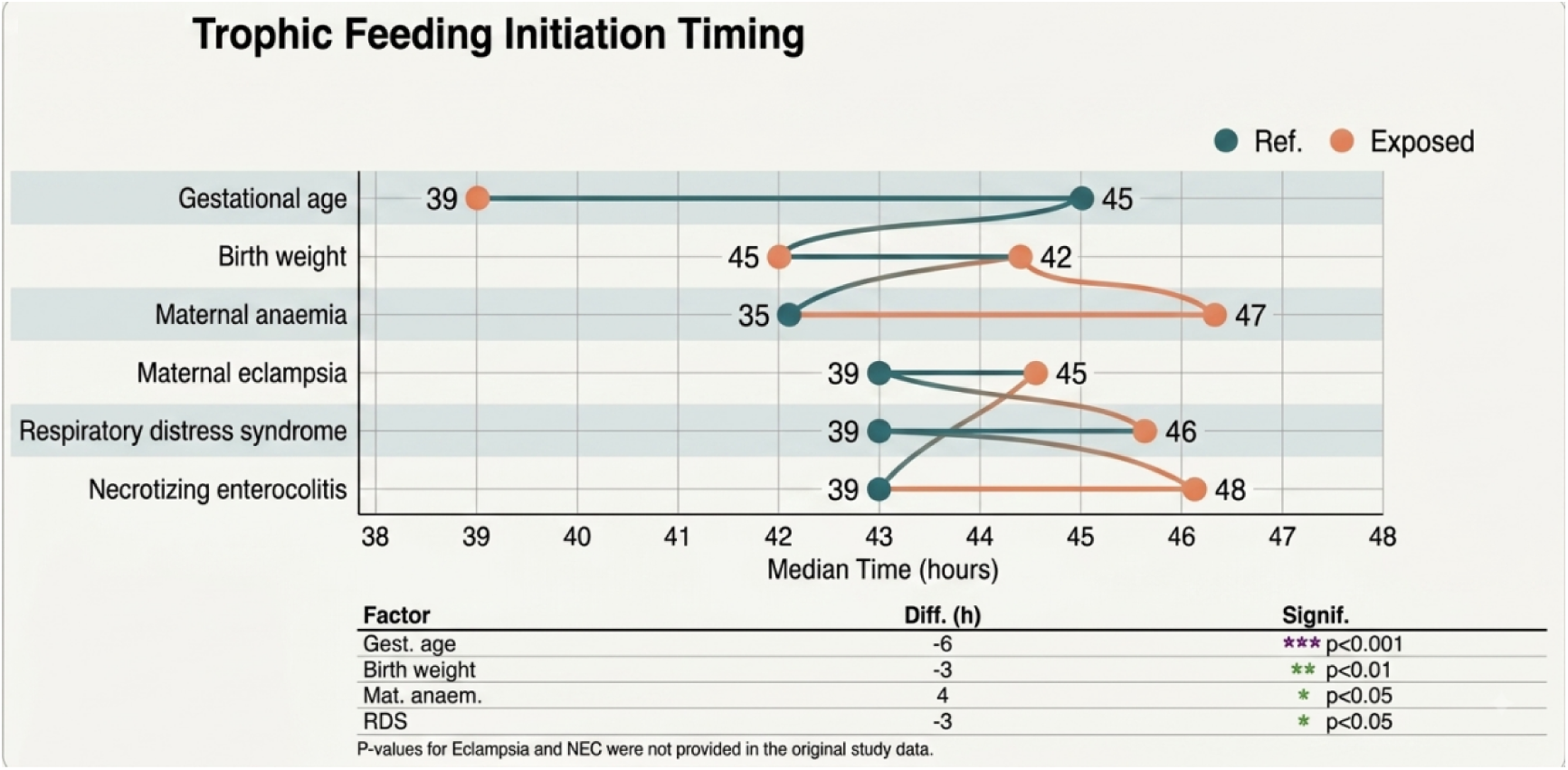
Trophic Feeding Initiation Timing. Median trophic feeding initiation time across clinical factors. Differences evaluated via log-rank tests (P < 0.05).

Gestational maturity was significantly associated with the timing of trophic feeding initiation. Neonates born at 34–36 weeks initiated feeding earlier than those born at 28–33 weeks, with median initiation times of 39 and 45 hours, respectively (log-rank P=0.0002). Similarly, neonates weighing ≥1500 g at birth initiated trophic feeding earlier than those weighing <1500 g, with median initiation times of 42 and 45 hours, respectively (log-rank P=0.020). The corresponding Kaplan–Meier curves showed a more rapid decline in the probability of remaining without feeding among more mature and heavier neonates, indicating earlier achievement of trophic feeding.

Maternal complications were also associated with feeding initiation time. Neonates born to mothers with anaemia had delayed initiation compared with those born to mothers without anaemia, with median times of 47 versus 39 hours, respectively (log-rank P=0.002). Maternal eclampsia was similarly associated with delayed feeding initiation, with a median time of 45 hours compared with 39 hours among neonates whose mothers did not have eclampsia.

Among neonatal clinical conditions, respiratory distress syndrome was significantly associated with delayed trophic feeding initiation. Neonates with respiratory distress syndrome initiated feeding at a median of 46 hours compared with 39 hours among those without the condition (log-rank P=0.004). Necrotizing enterocolitis showed an even stronger pattern of delayed initiation, with a median feeding initiation time of 48 hours among affected neonates compared with 39 hours among those without necrotizing enterocolitis. Collectively, these findings indicate that lower gestational maturity, lower birth weight, maternal complications, respiratory instability, and gastrointestinal morbidity were associated with prolonged time to trophic feeding initiation.

Figure 3. Kaplan–Meier estimates of time to trophic feeding initiation according to selected maternal and neonatal characteristics among preterm neonates. (A) Gestational age: 34–36 versus 28–33 weeks (log-rank P=0.0002); (B) birth weight: ≥1500 versus <1500 g (log-rank P=0.020); (C) maternal anaemia: yes versus no (log-rank P=0.002); (D) maternal eclampsia: yes versus no; (E) respiratory distress syndrome: yes versus no (log-rank P=0.004); and (F) necrotizing enterocolitis: yes versus no. Curves represent the probability of remaining without trophic feeding initiation over time; lower survival probabilities indicate earlier feeding initiation.

Figure 3. Kaplan–Meier curves for time to trophic feeding initiation according to selected maternal and neonatal characteristics

### 4. Predictors of time to trophic feeding initiation

Bivariable Cox proportional hazards regression was initially performed to identify variables potentially associated with time to trophic feeding initiation. Variables with P<0.25 in the bivariable analysis were considered for inclusion in the multivariable model. These included gestational age, birth weight, mode of delivery, first-minute APGAR score, maternal anaemia, maternal eclampsia, respiratory distress syndrome, perinatal asphyxia, and necrotizing enterocolitis.

After adjustment for potential confounding factors, gestational age, birth weight, maternal anaemia, respiratory distress syndrome, and necrotizing enterocolitis remained independently associated with time to trophic feeding initiation (Table 4; Figure 4).

**Figure 4.** Diagnostic assessment of the multivariable Cox proportional hazards model. Diagnostic assessment of the multivariable Cox proportional hazards model for time to trophic feeding initiation among preterm neonates admitted to the NICU at Adama Hospital Medical College, Southeast Ethiopia, 2024. (A) Schoenfeld residual assessment demonstrating no evidence of violation of the proportional hazards assumption (global test P=0.5253); and (B) Cox–Snell residual assessment showing close agreement between the estimated cumulative hazard and the 45-degree reference line, supporting adequate overall model fit.

**Figure 4a.**
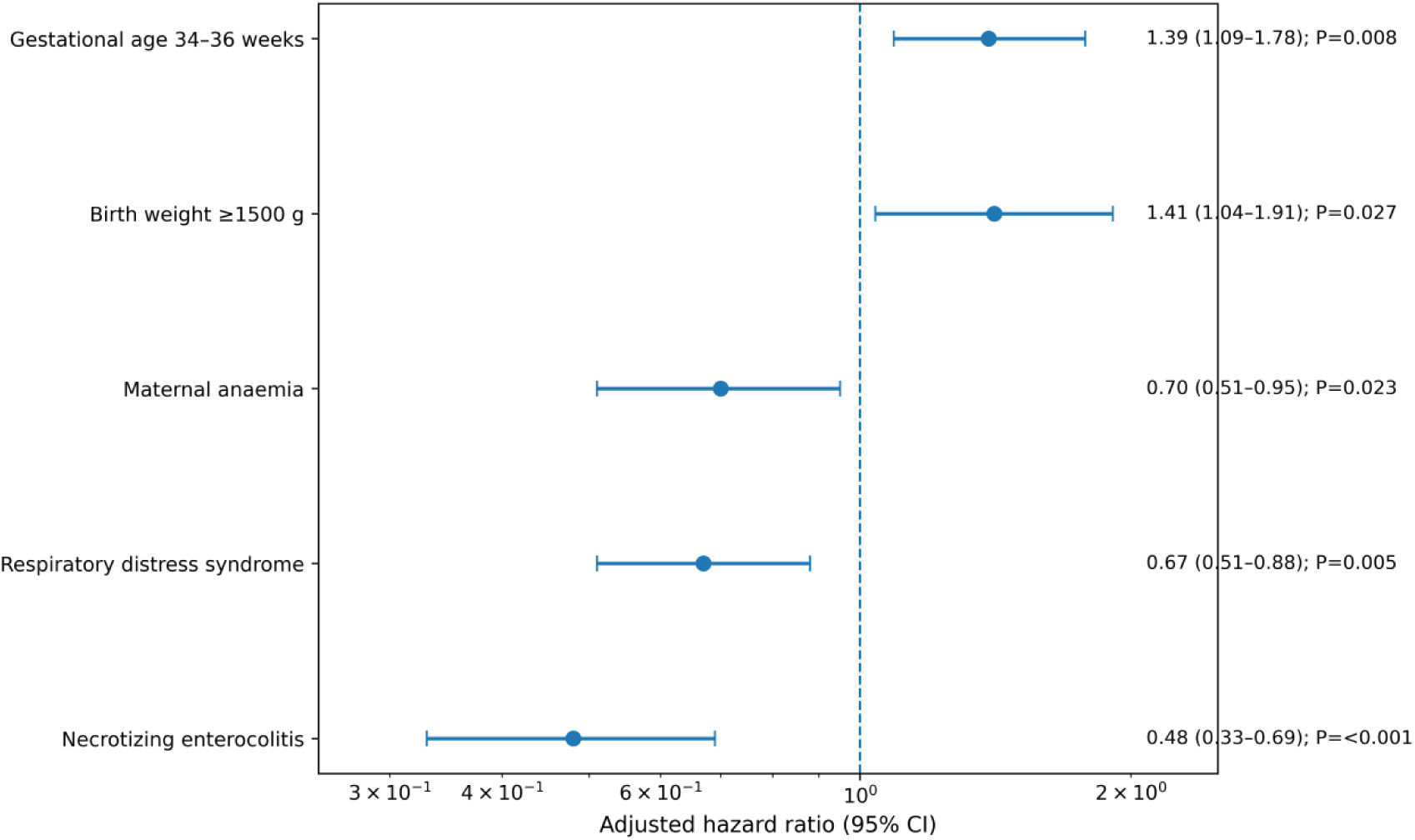
Forest plot of independent predictors of trophic feeding initiation. Forest plot showing adjusted hazard ratios for independent predictors of trophic feeding initiation among preterm neonates admitted to the NICU. The plot displays adjusted hazard ratios and 95% confidence intervals from the final multivariable Cox proportional hazards model. Gestational age of 34–36 weeks and birth weight ≥1500 g were associated with earlier trophic feeding initiation, whereas maternal anaemia, respiratory distress syndrome, and necrotizing enterocolitis were associated with delayed initiation after adjustment for potential confounding factors.

**Table 4.**
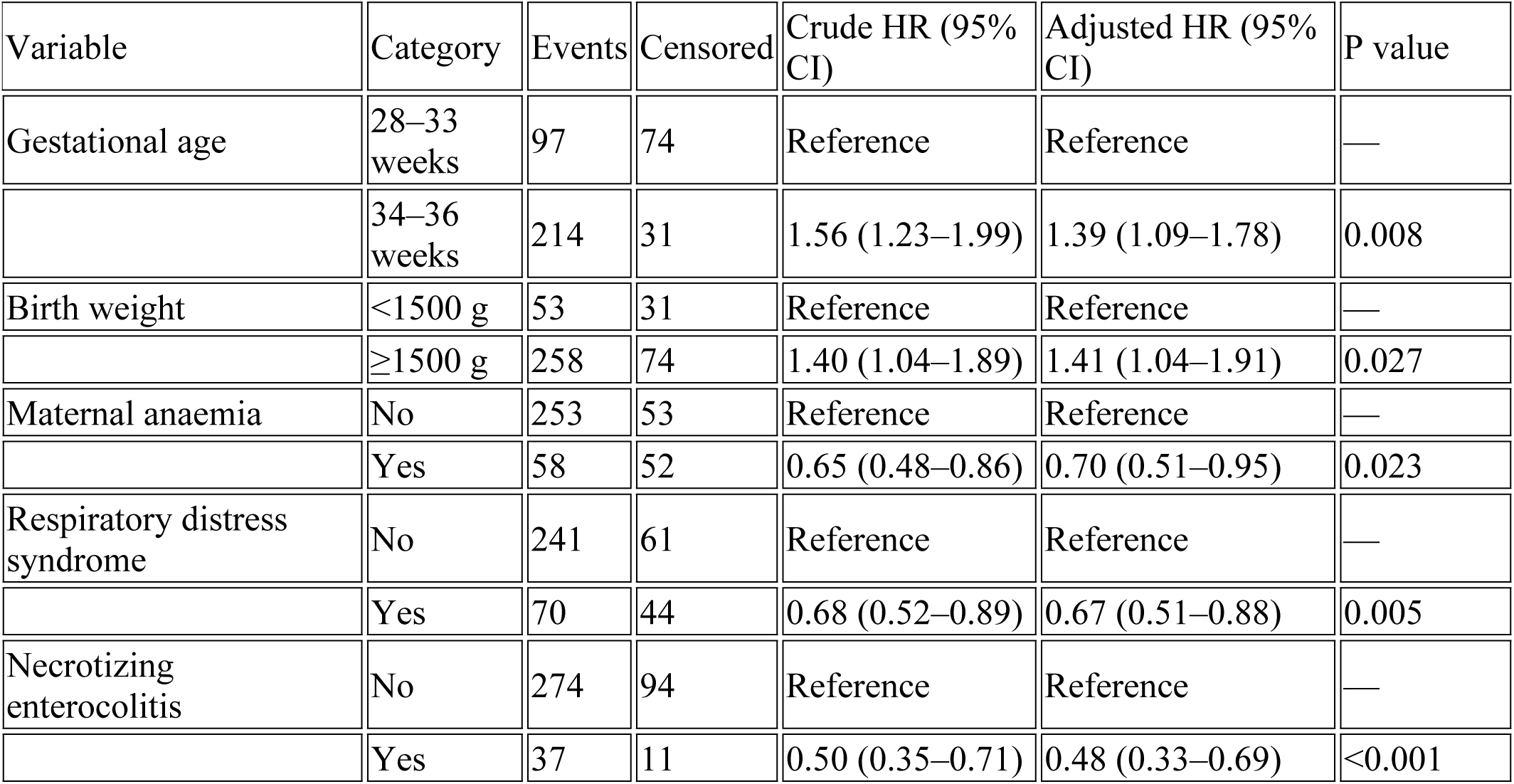
Bivariable and multivariable Cox proportional hazards regression analysis of predictors of trophic feeding initiation. presents crude and adjusted hazard ratios from Cox proportional hazards regression for time to trophic feeding initiation. Variables with P<0.25 in bivariable analysis were considered for the multivariable model. AHRs >1 indicate a higher rate and therefore earlier likelihood of trophic feeding initiation, whereas AHRs <1 indicate a lower rate and therefore delayed initiation. HR, hazard ratio; CI, confidence interval; AHR, adjusted hazard ratio.

Neonates born at 34–36 weeks of gestation had a significantly higher rate of trophic feeding initiation than those born at 28–33 weeks (AHR 1.39; 95% CI, 1.09–1.78; P=0.008), indicating a greater likelihood of initiating feeding earlier during follow-up. Similarly, neonates with a birth weight ≥1500 g had a higher rate of feeding initiation than those weighing <1500 g (AHR 1.41; 95% CI, 1.04–1.91; P=0.027).

In contrast, maternal and neonatal complications were associated with a lower rate of feeding initiation. Neonates born to mothers with anaemia had a significantly lower rate of trophic feeding initiation than those born to mothers without anaemia (AHR 0.70; 95% CI, 0.51–0.95; P=0.023). Respiratory distress syndrome was similarly associated with delayed feeding initiation (AHR 0.67; 95% CI, 0.51–0.88; P=0.005). Necrotizing enterocolitis demonstrated the strongest association with delayed feeding initiation; affected neonates had approximately half the rate of trophic feeding initiation compared with those without necrotizing enterocolitis (AHR 0.48; 95% CI, 0.33–0.69; P<0.001).

Although mode of delivery, first-minute APGAR score, maternal eclampsia, and perinatal asphyxia met the screening criterion for consideration in the multivariable analysis, their associations were attenuated after adjustment and they did not remain statistically significant in the final model. The complete bivariable and multivariable Cox regression results are presented in Table 4, while the magnitude and direction of the independent associations are visualized in Figure 5.

**Figure 5.**
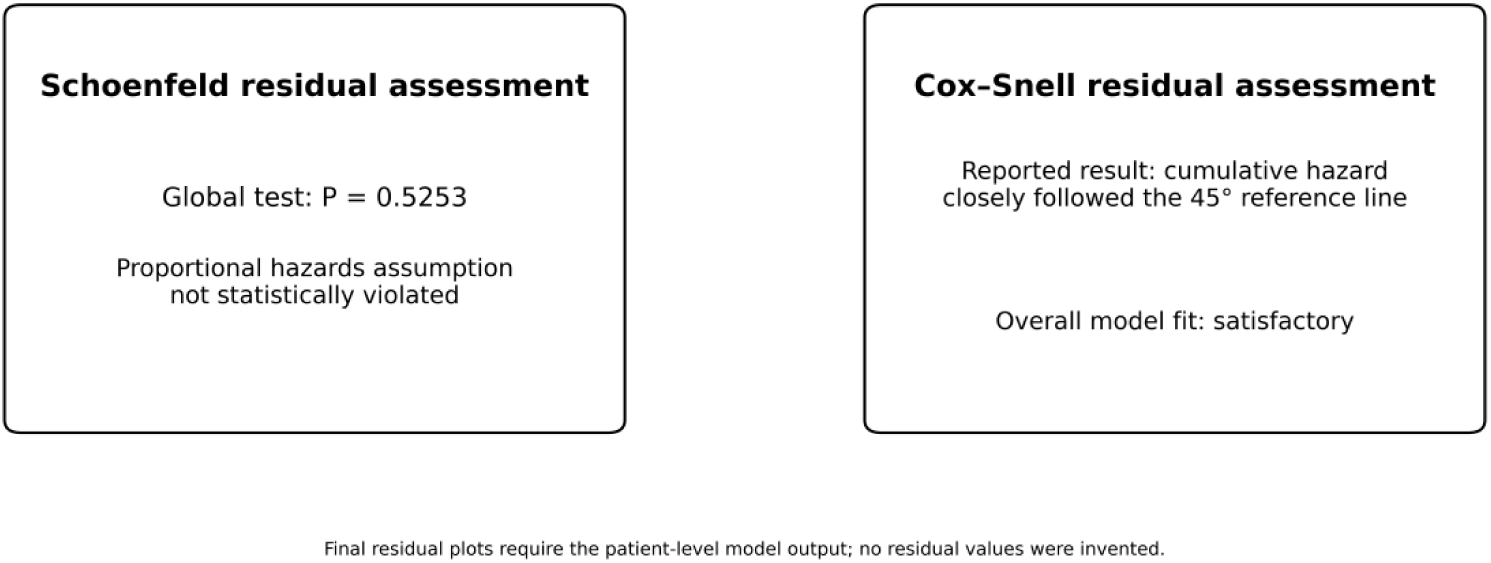
Diagnostic assessment of the multivariable Cox proportional hazards model.

### 6 Model diagnostics and goodness-of-fit assessment

The assumptions and adequacy of the final Cox proportional hazards model were evaluated before interpretation of the adjusted hazard ratios. The proportional hazards assumption was assessed using Schoenfeld residuals. The global test was not statistically significant (P=0.5253), providing no evidence of a violation of the proportional hazards assumption. Examination of the individual covariate residual patterns also showed no systematic departures over time, supporting the stability of the estimated hazard ratios.

Overall model adequacy was further evaluated using Cox–Snell residuals. The cumulative hazard of the residuals closely approximated the 45-degree reference line, indicating good agreement between the observed and expected survival experience and supporting satisfactory model fit. The diagnostic assessments are presented in (table 6 and Figure 5).

**Table 6.**
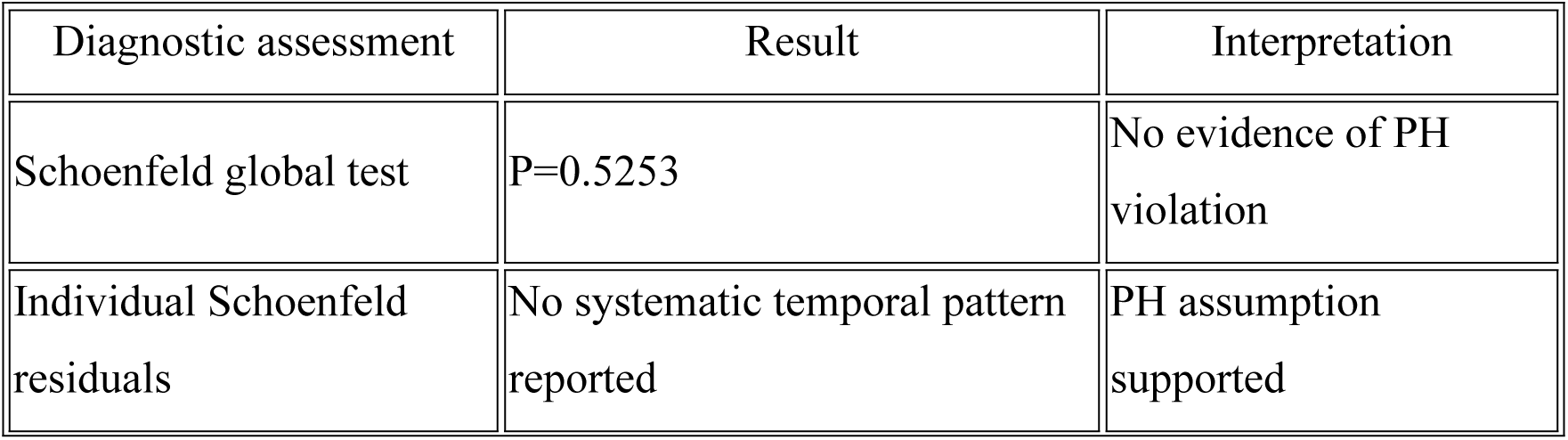
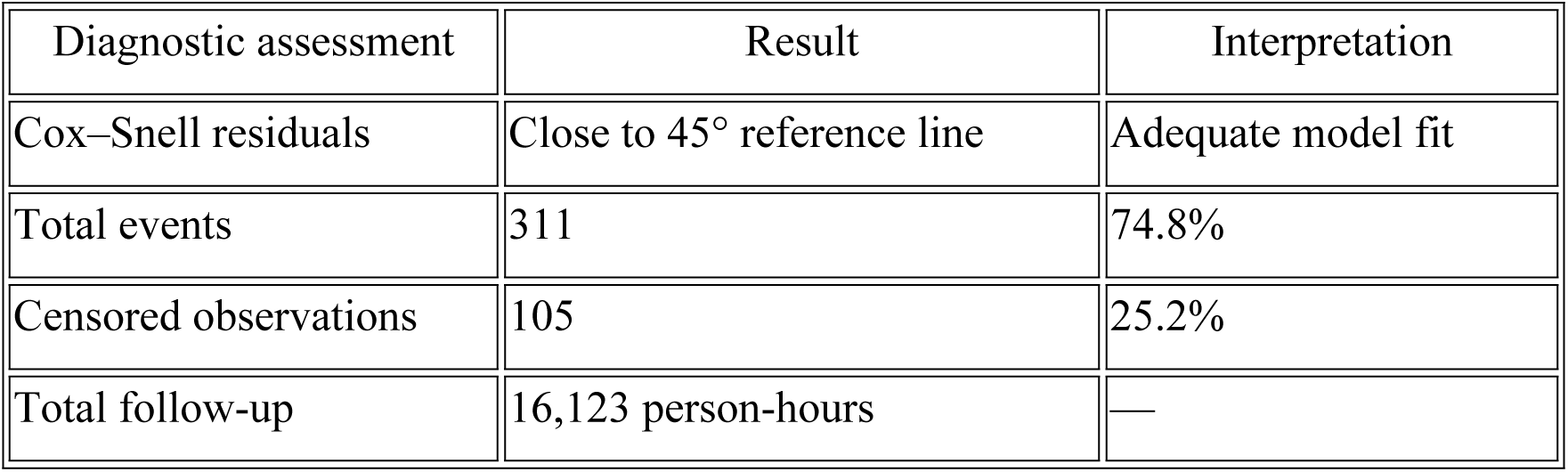
Model diagnostics. A compact diagnostic table will make the statistical reporting stronger without overloading the Results section. Diagnostics and adequacy of the final Cox proportional hazards model. The diagnostic assessments supported the validity and overall adequacy of the final multivariable model.

| Diagnostic assessment | Result | Interpretation |
| --- | --- | --- |
| Schoenfeld global test | $P=0.5253$ | No evidence of PH violation |
| Individual Schoenfeld residuals | No systematic temporal pattern reported | PH assumption supported |
| Cox–Snell residuals | Close to 45° reference line | Adequate model fit |
| Total events | 311 | 74.8% |
| Censored observations | 105 | 25.2% |
| Total follow-up | 16,123 person-hours | — |

### 7 Independent predictors of trophic feeding initiation

The adjusted analysis identified both neonatal maturity-related characteristics and clinical complications as independent determinants of the timing of trophic feeding initiation. Greater gestational age and higher birth weight were associated with earlier initiation, whereas maternal anaemia, respiratory distress syndrome, and necrotizing enterocolitis were associated with delayed initiation.

Among the independent predictors, necrotizing enterocolitis showed the largest reduction in the rate of feeding initiation (AHR 0.48; 95% CI, 0.33–0.69), followed by respiratory distress syndrome (AHR 0.67; 95% CI, 0.51–0.88) and maternal anaemia (AHR 0.70; 95% CI, 0.51–0.95). Conversely, neonates born at 34–36 weeks and those weighing ≥1500 g had higher rates of trophic feeding initiation than their respective reference groups. The direction, magnitude, and precision of these independent associations are summarized in the forest plot in Figure 5.

## 4. Discussions

The aim of this retrospective cohort study was to assess the timing of initiation of trophic feeding and its predictors among preterm neonates admitted to a neonatal intensive care unit in Southeast Ethiopia. The median time to start trophic feeding was 42 hours (IQR 24–50 hours) and approximately three-quarters of neonates started trophic feeding during follow-up. At the time of initiation of feeding, the practice was later than recommended international practices where early minimal enteral feeding within first 24 hours after birth is encouraged for clinically stable preterm infants. Early trophic feeding has been shown to improve gastrointestinal maturation, feeding tolerance, development of intestinal microbiota and better nutritional outcomes in very preterm and very low birth weight infants [5,9,11]. The observed delay underscores the persistent challenges in achieving evidence-based enteral nutrition practices in resource-limited NICU settings, where prematurity continues to be a major driver of neonatal morbidity and mortality [1–3].

The median time of initiation of feeding in this study was similar to previous studies done in Ethiopia [25, 26]. The median time of initiation of feeding was about 42 hours in Northern Ethiopia study [25] and 41 hours in Addis Ababa study [26]. The similarity could be due to the common clinical practice, neonatal care protocols, training of health care providers, and availability of resources for NICUs in Ethiopia. However, the delay beyond the first 24 hours is reflective of the fact that barriers to early trophic feeds still exist, despite the increasing recognition of the benefits of early trophic feeds. These barriers include neonatal instability, limited implementation of standardized feeding protocols, delayed clinical decision making and variation in provider practices. Neonatal nutrition studies have provided evidence that standardized feeding approaches can improve consistency of care and reduce unnecessary delays in enteral nutrition amongst preterm infants [17,30,32].

Initiation of trophic feeding was an important determinant of gestational age. Neonates who were born at 34-36 weeks of gestation started feeding earlier than those born at lower gestational ages. This finding is consistent with prior evidence that increased gestational maturity is associated with improved gastrointestinal development, improved coordination of sucking and swallowing reflexes and increased physiological stability, which promote earlier transition to enteral nutrition [11,15,16]. Very preterm infants, however, frequently have immature gastrointestinal function, respiratory instability and an increased risk of feeding intolerance or necrotizing enterocolitis, which often results in delaying the initiation of feeding until the infant is sufficiently stabilized [6,10].

Birth weight was also independently associated with earlier initiation of trophic feeding. Neonates ≥1500 g in weight started trophic feeding earlier than neonates <1500 g in weight. Similar associations have been reported in neonatal cohorts where birth weight is an important determinant of nutritional management and feeding tolerance [21,30]. Higher birth weight in infants is commonly associated with better physiologic reserves, lower risk of severe complications, and greater ability to tolerate enteral nutrition. Conversely, low birth weight infants are more prone to feeding intolerance, growth failure and necrotizing enterocolitis and therefore require close monitoring, cautious advancement of feeds and individualized nutritional strategies [32,33].

Maternal anaemia was associated with delayed initiation of trophic feeds. The finding highlights the impact of maternal nutritional status and pregnancy complications on early neonatal outcomes. Maternal anaemia has been linked to an increased risk of adverse pregnancy outcomes, including fetal growth restriction, complications of preterm birth and neonatal vulnerability, which may influence early feeding choices [25,26]. Infants of mothers with anaemia may require further clinical stabilization before initiation of enteral feeding is safe. Thus, strengthening of maternal nutrition interventions such as prevention and early screening and treatment of anaemia during pregnancy continues to be an important strategy to improve newborn survival and nutritional outcomes [25,27].

Another important predictor of delayed trophic feeding initiation was respiratory distress syndrome. Neonates who developed respiratory distress were fed later than those who did not have respiratory complications. This finding is consistent with previous studies showing that respiratory morbidity remains a major determinant of clinical instability and nutritional management of preterm infants [4,18,23]. Feeding may be a safety issue and a risk for aspiration in patients with respiratory distress that often requires oxygen therapy, continuous positive airway pressure, or mechanical ventilation. Moreover, respiratory compromise may impact gastrointestinal perfusion and motility and increase the risk of feeding intolerance. Thus, clinicians may defer the initiation of enteral feeding until respiratory status improves and feeding tolerance is expected to be established.

Delayed initiation of trophic feeding was the strongest factor associated with necrotizing enterocolitis. Neonates with necrotizing enterocolitis were significantly less likely to start trophic feeding than unaffected neonates. This finding is clinically plausible since necrotizing enterocolitis is a severe gastrointestinal inflammatory disease characterized by intestinal injury, feeding intolerance, abdominal distension and risk of intestinal perforation [10]. Standard neonatal management routinely holds enteral feeds during acute disease episodes to allow the bowel to recover and prevent progression. Prior evidence has suggested important roles of feeding strategies, human milk use and standardized feeding protocols in reducing NEC risk and improving feeding outcomes among very preterm infants [6–8,12,17].

In bivariable analysis, maternal eclampsia, perinatal asphyxia, mode of delivery and APGAR score were associated with trophic feeding initiation. However, these factors were not statistically significant after adjustment. This could mean that their effects are mediated by other clinical factors such as gestational age, birth weight, respiratory status and general neonatal condition. The findings emphasize the necessity of considering the combined effects of maternal, neonatal and clinical characteristics rather than individual factors in the design of feeding strategies for preterm infants.

The finding has important implications for neonatal care, public health programming and health system planning. At the facility level, unnecessary delays in the initiation of feeding can be reduced through standardized evidence-based trophic feeding protocols, regular clinical auditing, multidisciplinary neonatal nutrition teams and the promotion of human milk feeding [12,17,24]. There is a need to pay special attention to the vulnerable groups such as very pre-term babies, low birth weight babies and babies with respiratory or gastrointestinal problems. At the population level, improving maternal nutrition services, improving antenatal screening and treatment of maternal anemia and increasing access to quality obstetric and neonatal care may contribute to improved neonatal nutrition outcomes and survival [25–27].

These findings provide evidence for integration of early enteral feeding strategies in national neonatal care guidelines for policy makers and health planners, strengthening of NICU infrastructure, improving availability of human milk support services and ensuring adequate training of neonatal healthcare providers . These interventions align with global newborn survival priorities to reduce preventable neonatal morbidity and mortality, particularly in low-resource settings where prematurity and neonatal complications remain major public health challenges [1,25,27].

Future research should consider multicentre prospective cohorts from different levels of neonatal care facilities to improve generalisability of findings. Additional studies should explore practices of healthcare providers, barriers to protocol implementation, maternal breast milk availability, patterns of feeding intolerance, and long-term growth and neurodevelopmental outcomes associated with timing of initiation of trophic feeding. Interventional studies are required to test standardized feeding protocols, quality improvement strategies, and context-specific neonatal nutrition strategies to optimize early enteral nutrition in preterm neonates in low-resource settings [32,33].

Strength and Limitation

Strength

Limitation

Conclusion

Recommendation

## Data Availability

No datasets were generated or analysed during the current study.

## Declaration

### Ethics approval and consent to participate

Ethical clearance was obtained from the Institutional Review Board of Adama Hospital Medical College (Protocol number: 0147/K-373/2024). Permission for the study was obtained from the hospital administration and Neonatal Intensive Care Unit. As this retrospective cohort study used anonymised routinely collected neonatal medical record data, individual informed consent was waived by the ethics committee. All data were handled with confidentiality and the study was carried out in accordance with the Declaration of Helsinki.

### Consent to publish

N/A. The study utilized anonymised secondary data from neonatal medical records and did not include any information that could identify a single individual.

### Data and materials availability

The dataset generated and analysed during this study is not publicly available due to confidentiality requirements of neonatal clinical records. An anonymised dataset is available from the corresponding author upon reasonable request and with permission from the relevant institutional authorities.

### Authors contributions

BM conceived and designed the study, developed the protocol, coordinated and supervised data collection, contributed to data analysis and interpretation and drafted the manuscript. GA was involved in implementation of the study, data collection, data management, interpretation of findings and critical revision of the manuscript. IM helped to collect data, do statistical analysis, interpret results and revise the manuscript. All authors have contributed to the critical revision of the manuscript, have approved the final version and agree to be accountable for all aspects of the work.

## Acknowledgments

We would like to thank Adama Hospital Medical College, Neonatal Intensive Care Unit staff, medical record personnel, data collectors and supervisors for their valuable support during the study. The authors would also like to thank all the health care professionals who were involved in the management of preterm neonates and whose clinical records contributed to this research.

